# *Plasmodium falciparum pfhrp2* and *pfhrp3* gene deletions among patients enrolled at 100 health facilities throughout Tanzania: February to July 2021

**DOI:** 10.1101/2023.07.29.23293322

**Authors:** Eric Rogier, Nastassia Battle, Catherine Bakari, Misago D. Seth, Douglas Nace, Camelia Herman, Rashid A. Madebe, Celine I. Mandara, Beatus M. Lyimo, David J. Giesbrecht, Zachary R. Popkin-Hall, Filbert Francis, Daniel Mbwambo, Issa Garimo, Sijenunu Aaron, Abdallah Lusasi, Fabrizio Molteni, Ritha Njau, Jane A. Cunningham, Samwel Lazaro, Ally Mohamed, Jonathan J. Juliano, Jeffrey A. Bailey, Venkatachalam Udhayakumar, Deus S. Ishengoma

**Affiliations:** Malaria Branch, Centers for Disease Control and Prevention, Atlanta, GA, USA; CDC Foundation, Atlanta, GA; National Institute for Medical Research, Dar es Salaam, Tanzania; Nelson Mandela African Institution of Science and Technology, Arusha, Tanzania; Brown University, Providence, MA, USA; University of North Carolina, Chapel Hill, NC, USA; National Institute for Medical Research, Tanga, Tanzania; National Malaria Control Programme, Dodoma, Tanzania; Swiss Tropical Public Health Institute, Dar es Salaam, Tanzania; World Health Organization, Country Office, Dar es Salaam, Tanzania; World Health Organization, Headquarters, Geneva, Switzerland; Faculty of Pharmaceutical Sciences, Monash University, Melbourne, Australia; Harvard T.H Chan School of Public Health, Boston, MA, USA

**Author notes:** Correspondence: Eric Rogier, Centers for Disease Control and Prevention, Atlanta, GA 30029.

**Keywords:** *Plasmodium falciparum*, Tanzania, RDT, *pfhrp2*, *pfhrp3*, gene deletion

## Abstract

*Plasmodium falciparum* with the histidine rich protein 2 gene (*pfhrp2*) deleted from its genome can escape diagnosis by HRP2-based rapid diagnostic tests (HRP2-RDTs). The World Health Organization (WHO) recommends switching to a non-HRP2 RDT for *P. falciparum* clinical case diagnosis when *pfhrp2* deletion prevalence causes ≥ 5% of RDTs to return false negative results. Tanzania is a country of heterogenous *P. falciparum* transmission, with some regions approaching elimination and others at varying levels of control. In concordance with the current recommended WHO *pfhrp2* deletion surveillance strategy, 100 health facilities encompassing 10 regions of Tanzania enrolled malaria-suspected patients between February and July 2021. Of 7,863 persons of all ages enrolled and providing RDT result and blood sample, 3,777 (48.0%) were positive by the national RDT testing for *Plasmodium* lactate dehydrogenase (pLDH) and/or HRP2. A second RDT testing specifically for the *P. falciparum* LDH (Pf-pLDH) antigen found 95 persons (2.5% of all RDT positives) were positive, though negative by the national RDT for HRP2, and were selected for *pfhrp2* and *pfhrp3* (*pfhrp2/3*) genotyping. Multiplex antigen detection by laboratory bead assay found 135/7,847 (1.7%) of all blood samples positive for Plasmodium antigens but very low or no HRP2, and these were selected for genotyping as well. Of the samples selected for genotyping based on RDT or laboratory multiplex result, 158 were P. falciparum DNA positive, and 140 had sufficient DNA to be genotyped for pfhrp2/3. Most of these (125/140) were found to be pfhrp2+/pfhrp3+, with smaller numbers deleted for only pfhrp2 (n=9) or only pfhrp3 (n=6). No dual pfhrp2/3 deleted parasites were observed. This survey estimated that 0.24% (95% confidence interval: 0.08% to 0.39%) of false-negative HRP2-RDTs for symptomatic persons were due to pfhrp2 deletions in this 2021 Tanzania survey. These data provide evidence for HRP2-based diagnostics as currently accurate for P. falciparum diagnosis in Tanzania.

## Introduction

Scaling-up of malaria control interventions since the year 2000 has resulted in dramatic decreases in *Plasmodium falciparum* cases and deaths in sub-Saharan Africa [1, 2]. However, due to disruption of services in the years 2020 and 2021 due to the SARS-CoV-2 pandemic, an estimated 50,000 additional malaria deaths occurred during each of those years [1], showing how gains from malaria control can quickly be lost. Malaria control efforts can take many different forms, including (but not limited to) insecticide treated net distribution, accurate parasitological confirmation of *Plasmodium* infections, and effective case management by provision of efficacious antimalarial drugs. Even these pillars of malaria control are being threatened in sub-Saharan Africa through vector insecticide resistance [3, 4], *P. falciparum* diagnostic resistance by *pfhrp2* gene deletions [5–7], and emergence of antimalarial drug resistance genotypes [8].

Use of antigen-detecting rapid diagnostic tests (RDTs) has been a pragmatic strategy for parasitological confirmation of *P. falciparum* infection through detection of *Plasmodium* antigens, and has enabled better targeting of treatment and improved surveillance of malaria [9]. The histidine-rich protein 2 (HRP2) antigen is a species-specific target for *P. falciparum*, though detection of the *Plasmodium* lactate dehydrogenase (LDH) antigen can also be a reliable diagnostic marker – especially for higher parasite density infections typically seen in clinical cases. RDT products have been developed which have capture/detection antibodies which would bind any *Plasmodium* LDH (pan-*Plasmodium*), or the *P. falciparum-* and *P. vivax-* specific epitopes of this antigen [10]. Similar to the HRP2 antigen, HRP3 is expressed by *P. falciparum* and can also supplement the detection signal for HRP2-based RDTs or other immunoassays [9, 11]. Given the predominance of *P. falciparum* for the clinical relevance and malaria epidemiology in sub-Saharan Africa, HRP2-based RDTs are highly preferred because of thermal stability, abundance of antigen target, high species specificity, and lower cost [12].

Deletions of the *pfhrp2* and *pfhrp3* (*pfhrp2/3*) genes from the *P. falciparum* genome have now been observed in multiple African countries and pose a serious threat to the use of these antigenic markers to reliably detect *P. falciparum* infection [5–7, 9]. Natural parasite populations with *pfhrp2/3* gene deletions were first reported in Peru in 2010 [13], but *P. falciparum* has since been found with these gene deletions in many settings [6, 14], and has now been reported in more than 35 countries [15]. The World Health Organization (WHO) has issued guidance on investigating suspected false-negative RDT results due to *pfhrp2/3* gene deletions in malaria parasites and is encouraging a harmonised approach to mapping and reporting the deletions [16, 17]. Based on these guidelines, each country should endeavour to establish the status and monitor gene deletion trends over time to see if re-assessment of national diagnostic strategy is warranted when ≥5% of *P. falciparum* clinical infections are missed by HRP2-RDTs. In Tanzania, previous reports have shown sporadic occurrence of *pfhrp2* gene deletions in multiple locations, though at very low levels (<2.0%) [18, 19]. However, all Tanzanian studies reported to-date have utilized sampling designs different from WHO recommendations for reporting of nationwide gene deletion estimates. This current study provides the first national evaluation of *pfhrp2/3* gene deletions in the *P. falciparum* parasite population and provides baseline data for future monitoring to establish the temporal and spatial trends of these deletions in the country.

## Methods

### Ethics

The study protocol was adopted from the WHO template [17] and submitted to the Tanzanian Medical Research Coordinating Committee (MRCC) of the National Institute for Medical Research (NIMR) for review and ethical approval. The protocol was also submitted for review and approval by the ethics committee of WHO in Geneva, Switzerland. All research participants were asked and provided individual consent (or assent for children aged 7-18 years of age) for their participation in the survey and biobanking for future research. For children under the legal age of adulthood in Tanzania (<18 years), consent was obtained from a parent or guardian. An informed consent form was developed in English and translated in Kiswahili and used to obtain consent both verbally and in writing from all participants. All participants agreed and signed the consent or assent form or provided a thumbprint in conjunction with the signature of an independent witness in case the study participant was illiterate. The project was approved as a program evaluation activity and non-human subjects research by the U.S. Centers for Disease Control and Prevention (CDC, project ID: 0900f3eb81f722fb).

### Study design

The cross-sectional study which was undertaken as part of the ongoing project on the molecular surveillance of malaria in Tanzania (MSMT)[20], conducted in collaboration with the Tanzanian National Malaria Control Programme (NMCP), the President’s Office, Regional Administration, and Local Government authority (PO-RALG) and collaborators from USA (Brown University, University of North Carolina and the CDC). Ten regions of Tanzania were utilized and 100 health facilities (HF) were selected according to WHO protocol as enrolment sites [21]: Dar es salaam, Dodoma, Kagera, Kilimanjaro, Manyara, Mara, Mtwara, Njombe, Songwe and Tabora (Figure 1). At each HF (10 HFs from each region), patients presenting with malaria-like illness were approached and to enrolled in this study after providing an informed consent. Regardless of enrolment in this study, patients with malaria-like illness were provided an RDT, and if positive, treated with appropriate antimalarial drugs according to Tanzania national guidelines.

**Figure 1.**
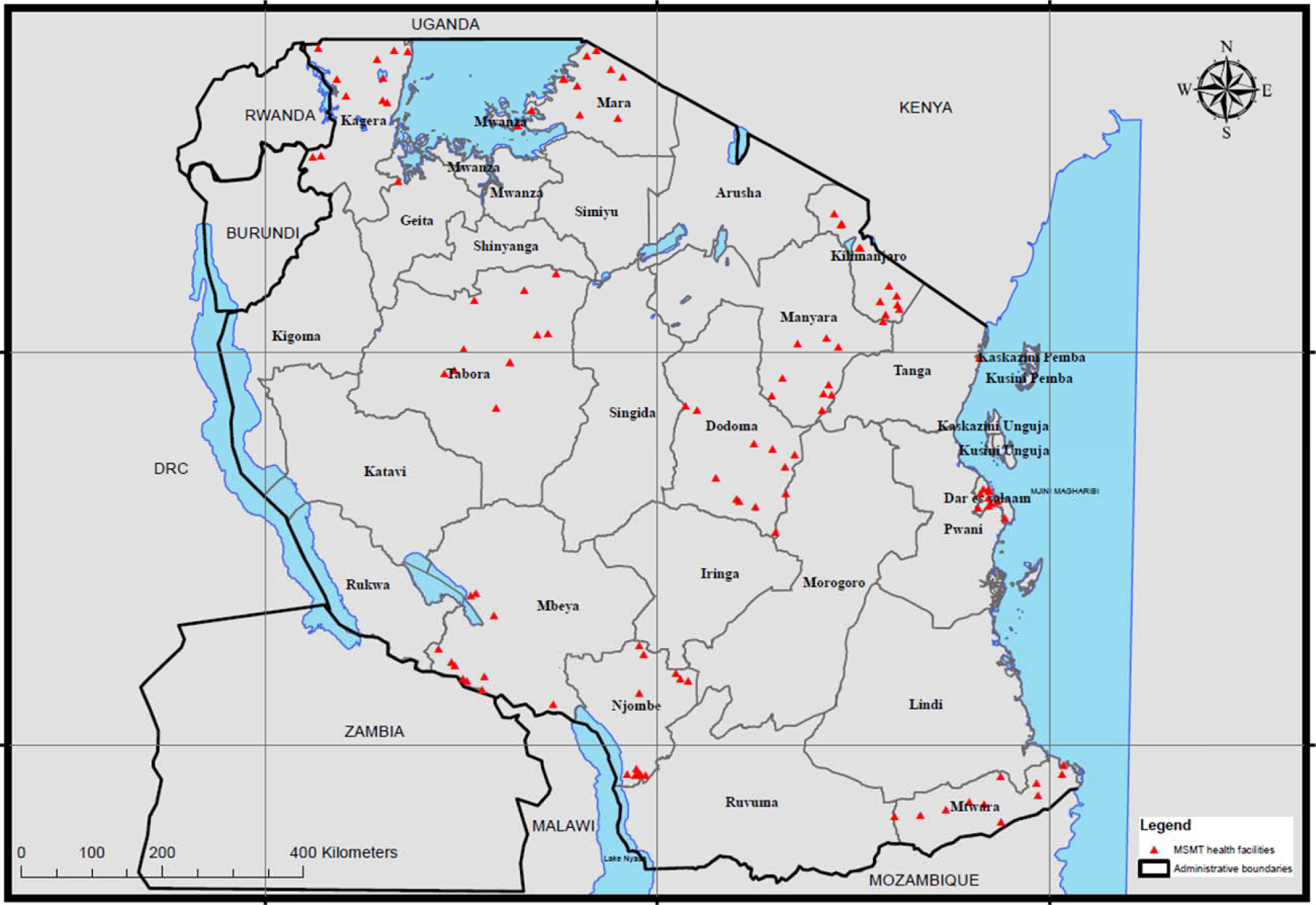
Map of showing health facility enrollment sites in 10 regions which were involved in the survey: Tanzania 2021. Individual health facilities are shown by red triangles.

### Patient enrollment and sample collection procedures

Inclusion criteria required patients to be aged 6 months and above, meeting case definition for suspected malaria (fever in the last 48 hrs or fever at presentation with axillary temperature ≥37.5°C and/or other malaria symptoms), and requirement to reside in the same area as the health facility. Exclusion criteria included previous enrollment in the study, severe malaria, or other severe illnesses. For each patient, finger prick blood was collected for detection of malaria infection by RDTs and also for preparation of dried blood spots (DBS) on Whatman 3 MM CHR filter papers (Cytiva, USA).

At enrollment, each patient was tested by two types of RDTs, with the first being the HRP2/pan-LDH test used by the Tanzanian NMCP for routine diagnosis of malaria (“national RDT”, listed below) and the second was the *P. falciparum* LDH test donated by WHO for this survey (“research RDT”), the #C14RHG25, RapiGEN Inc. BIOCREDIT Malaria Ag Pf (Pf-pLDH), Republic of Korea). At the time of the survey, the Tanzanian NMCP had three types of RDTs which were concurrently used in the country. They included SD Bioline Malaria Ag P.f/pan (#05FK60, Standard Diagnostic Inc., India), CareStart Malaria HRP2/pLDH (#RMOM-02571, AccessBio Inc., NJ, USA) and First Response Malaria Ag HRP2/pLDH Combo (#PI16FRC10s, Premier Medical Corp. India). “RDT discordance” for this study was defined as a positive Pf-pLDH result, but a negative HRP2 band.

Sample size for this survey was based on WHO master protocol [21], with a simple random sampling design effect of 1.5 accounting for potential deletions of the *pfhrp2* gene. A target of 37 patients with positive results by either of the two tests were enrolled from each HF for a total target size of 370 samples per region and 3,700 RDT positive persons for the whole study [21]. All DBS samples (positive and negative by RDTs) were stored at the HFs at ambient temperature and later shipped to the NIMR Genomics Laboratory after the end of the survey for storage at room temperature before shipping to the CDC in Atlanta, GA, USA for further analysis.

### Sample processing and multiplex bead assay

To rehydrate blood samples for *Plasmodium* antigen detection, a 6mm filter paper punch from each DBS (approximately 10 μL whole blood) was placed into 200 μL of elution buffer: phosphate buffered saline (PBS, Sigma) pH 7.2, 0.3% Tween-20, 0.5% casein (ThermoFisher), 0.5% bovine serum albumin (BSA, Sigma), 0.5% polyvinyl alcohol (Sigma), 0.8% polyvinlypyrrolidine (Sigma), 0.02% sodium azide (Sigma), and 3 μg/mL of *E. coli* lysate (to prevent nonspecific binding). This provided a 1:20 dilution of whole blood which was used as the test sample for the bead-based multiplex immunoassay.

The bead-based multiplex assay for malaria antigen detection was performed as described previously [22]. Magnetic microbeads (xMAP®, Luminex Corp., Austin, TX) were covalently bound to antibodies for *Plasmodium* antigen capture by the Luminex antibody coupling kit according to manufacturer’s instructions. Per 12.5x10^6^ beads, antibody coupling concentrations were: anti-pan-*Plasmodium* aldolase antibody (pAldolase, 12.5 μg, Abcam, Cambridge, UK); anti-pan-*Plasmodium* LDH (pLDH, 12.5 μg, clone M1209063, Fitzgerald, Acton, MA); anti-*P. falciparum* HRP2 (20 μg, clone MPFG-55A, ICLlabs, Portland, OR); anti-*P. falciparum* LDH (20 μg, clone M1209062, Fitzgerald). Detection antibodies were also prepared in advance by biotinylating (EZ-link Micro Sulfo-NHS-Biotinylation Kit, ThermoScientific, Waltham, MA) according to manufacturer’s instructions. Final prepared dilution of detection antibodies was 1.0 mg/mL and for anti-malarial antigen specific antibodies as follows: pAldolase (Abcam), LDH (1:1 antibody mixture of M1709Pv1 and M86550, Fitzgerald), HRP2 (1:1 antibody mixture of MPFG-55A and MPFM-55A, ICLlabs). All reagents were stored at 4°C until use in the immunoassay.

For the antigen multiplex assay, reagents were diluted in buffer containing 0.1M PBS (pH 7.2), 0.05% Tween-20, 0.5% BSA, and 0.02% sodium azide. Assay plates were affixed to a handheld magnet (Luminex Corp) for wash steps and allowed one minute for bead magnetization before evacuation of liquid and washing with 100 μL PBS/0.05% Tween-20. The four bead regions were combined in dilution buffer (in a reagent trough) and pipetted onto a 96- well assay plate (BioPlex Pro, BioRad, Hercules, CA) at a quantity of approximately 800 beads/region. Plates were washed twice, and 50 μL of controls (in duplicate) or samples (in singlet) were pipetted into appropriate wells. Following 90-minute gentle shaking at room temperature, protected from light, plates were washed three times. A mixture of detection antibodies was prepared in dilution buffer (pAldolase at 1:2000, all others at 1:500) and 50 μL added to each well for a 45-minute incubation. After three washes, 50 μL of streptavidin-phycoerythrin (at 1:200, Invitrogen) was added for a 30-minute incubation. Plates were washed three times, and 50 μL dilution buffer was added to each well for 30-minute incubation. Plates were then washed once and beads resuspended in 100 μL PBS. After brief shaking, plates were read on a MAGPIX machine (Luminex Corp) with a target of 50 beads per region. The median fluorescence intensity (MFI) value was generated for all beads collected for each region by assay well. Subtraction of the assay signal from wells with dilution buffer blank provided the MFI-background (MFI-bg) value used for analyses. Positive and negative controls were included on each assay plate to ensure valid assay results

### Selection of samples for PET-PCR Plasmodium speciation and DNA quality control

Samples were initially selected for PET-PCR speciation and *pfhrp2/3* genotyping if they had discordant results (point-of-care RDTs showed positivity for the Pf-pLDH band but a negative HRP2 band). Based on laboratory multiplex antigen data, the second selection of samples for Plasmodium speciation by PET-PCR and *pfhrp2/3* genotyping was done based on the relationship between the two pan-*Plasmodium* antigens (aldolase and LDH) and the HRP2 and/or HRP3 (HRP2/3) signal as described previously [22], or relationship between Pf-pLDH and HRP2/3. Samples were selected if completely lacking an assay signal for HRP2/3 or if the assay signal for HRP2/3 was atypically lower compared to the level of pAldolase, pLDH, or Pf-pLDH antigens.

### DNA extraction and quantification from samples

For samples selected for genotyping, total genomic DNA was extracted from DBS samples (three 6mm punches per sample) using Tween-Chelex 100 (Bio-Rad Laboratories, USA) extraction method as previously described with some modifications [23]. Briefly, the punched DBS samples were incubated in 1 mL of 0.5% Tween-20 (Sigma) in PBS (Thermo Fisher Scientific) overnight at room temperature, placed onto the shaker at low speed to release parasite DNA from red blood cells, and briefly centrifuged. Tween-PBS was removed followed by washing with 1 mL of 1X PBS and incubation at 40°C for 30 min. The samples were briefly centrifuged, PBS removed, and 150 μL of 10% Chelex 100 resin (BioRad) in water were added to each sample and incubated for 10 min at 95°C. Supernatant was removed from the tubes followed by addition of 1 mL PBS, briefly vortexed, and incubated at 4°C for 15-30 min. After a brief centrifugation to precipitate the DNA from other impurities, a 2:1 solution of Chelex to sterile water was added and incubated at 95°C for 10 min. Samples were centrifuged for 5 min at high speed to pellet Chelex and filter paper and resulting DNA supernatants (approximately 150 μL) were transferred to new tubes. Supernatants were centrifuged for 10 min at 14,000 rpm, and the Chelex-free supernatant (approximately 50 μL) was transferred to a new tube for storage at 20°C until use in PCR assays.

### PET-PCR and genotyping for *P. falciparum pfhrp2* and *pfhrp3* genes

PET-PCR was performed on extracted DNA as described previously to ensure quality and presence of *P. falciparum* DNA [22]. Genotyping for *pfmsp1*, *pfmsp2*, and *pfhrp3* was performed by nested PCR (nPCR) as described previously [22]. For *pfhrp2* genotyping, conventional PCR was performed as described previously [24]. Results for *pfhrp2/3* genotyping were only reported if both *pfmsp1* and *pfmsp2* (both single-copy genes in the *P. falciparum* genome) were successfully amplified for a DNA sample in at least two out of three independent PCR reactions [16].

## Results

From February to July 2021, a total of 8,040 persons were enrolled among the 100 Tanzanian HFs (Figure 1) with a recorded RDT result and 7,863 persons (97.8%) providing DBS for later laboratory assays. Of this set of DBS, 7,847 (99.8%) were able to be inventoried and had laboratory multiplex antigen data collected. For all enrolled participants, median age was 15 (interquartile range: 3 to 29 years), and 55.3% were female (Table 1). Numbers of participants enrolled among regions were similar except Kilimanjaro which enrolled 31.6% of all patients; this was due to the very low transmission setting in Kilimanjaro region and need to enroll more persons to obtain the target of 37 RDT positive persons per health facility [21]. RDT results found 48.2% (n=3,787) of all patients positive by any RDT utilized in the survey. For the 3,787 patients who showed a positive RDT result to any antigen target, 3,185 (84.1%) were positive to both targets on the national RDT and 545 (14.4%) were positive only for the HRP2 band. The research RDT, testing only for Pf-pLDH, showed strong concordance with national RDT results and 3,020 (79.7%) of persons positive by the research RDT also showed positivity by the national RDT. Of these 767 testing positive only by the national RDT, and not the research RDT, the majority of these (n=609, 79.4%) were only HRP2 positive, indicating a lower-density *P. falciparum* infection [25]. A total of 95 persons tested positive only by the research RDT, representing discordance of 2.5% of the “any RDT” positives.

**Table 1.**
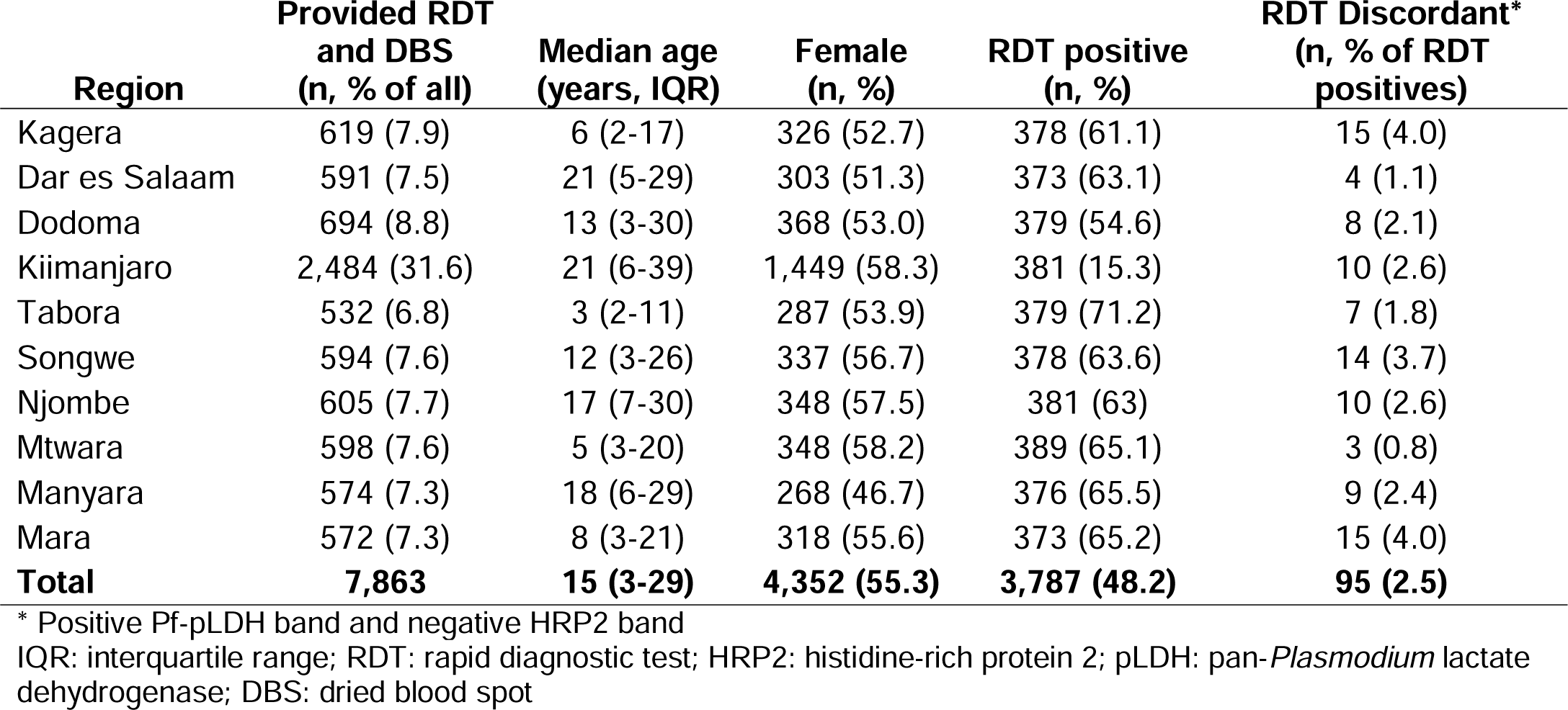
Demographic and RDT characteristics of enrolled patients: Tanzania 2021.

| Region | Provided RDT<br>and DBS<br>(n, % of all) | Median age<br>(years, IQR) | Female<br>(n, %) | RDT positive<br>(n, %) | RDT Discordant*<br>(n, % of RDT<br>positives) |
| --- | --- | --- | --- | --- | --- |
| Kagera | 619 (7.9) | 6 (2-17) | 326 (52.7) | 378 (61.1) | 15 (4.0) |
| Dar es Salaam | 591 (7.5) | 21 (5-29) | 303 (51.3) | 373 (63.1) | 4 (1.1) |
| Dodoma | 694 (8.8) | 13 (3-30) | 368 (53.0) | 379 (54.6) | 8 (2.1) |
| Kiimanjaro | 2,484 (31.6) | 21 (6-39) | 1,449 (58.3) | 381 (15.3) | 10 (2.6) |
| Tabora | 532 (6.8) | 3 (2-11) | 287 (53.9) | 379 (71.2) | 7 (1.8) |
| Songwe | 594 (7.6) | 12 (3-26) | 337 (56.7) | 378 (63.6) | 14 (3.7) |
| Njombe | 605 (7.7) | 17 (7-30) | 348 (57.5) | 381 (63) | 10 (2.6) |
| Mtwara | 598 (7.6) | 5 (3-20) | 348 (58.2) | 389 (65.1) | 3 (0.8) |
| Manyara | 574 (7.3) | 18 (6-29) | 268 (46.7) | 376 (65.5) | 9 (2.4) |
| Mara | 572 (7.3) | 8 (3-21) | 318 (55.6) | 373 (65.2) | 15 (4.0) |
| **Total** | **7,863** | **15 (3-29)** | **4,352 (55.3)** | **3,787 (48.2)** | **95 (2.5)** |

\* Positive Pf-pLDH band and negative HRP2 band
IQR: interquartile range; RDT: rapid diagnostic test; HRP2: histidine-rich protein 2; pLDH: pan-*Plasmodium* lactate dehydrogenase; DBS: dried blood spot

The DBS from these 95 persons testing positive by only Pf-pLDH were selected for further PCR assays based on suspicion of *P. falciparum* infection with potential deletions in the *pfhrp2* (and *pfhrp3*) genes. Of these 95, nine DBS (9.5%) were unable to be retrieved, and an additional 46 (48.4%) were negative for *P. falciparum* DNA, leaving 40 (42.1%) appropriate for further *pfhrp2/3* genotyping. Additionally, DBS were also selected for further PCRs based on results from the laboratory multiplex antigen detection assay. In comparing the laboratory-detected antigen levels of HRP2/3 versus other *Plasmodium* markers (assay signal for pAldolase, pLDH, and Pf-pLDH), if a blood sample was positive for any *Plasmodium* antigen, the vast majority of these were observed to have high levels of HRP2/3 relative to the other three targets (Figure 2). From the lab multiplex antigen assay, a total of 135 specimens (1.7% of the 7,847 DBS available; shown by red shading in Figure 2) were selected for DNA extraction and further assays to determine *pfhrp2/3* status of the infecting parasites. Of these 135, 118 (87.4%) were confirmed to have *P. falciparum* DNA to attempt the *pfhrp2/3* genotyping PCRs (Figure 3). Concordance of lab antigen positivity between the blood samples from the 95 persons testing RDT positive only for Pf-pLDH is shown in Supplemental Figure 1.

**Figure 2.**
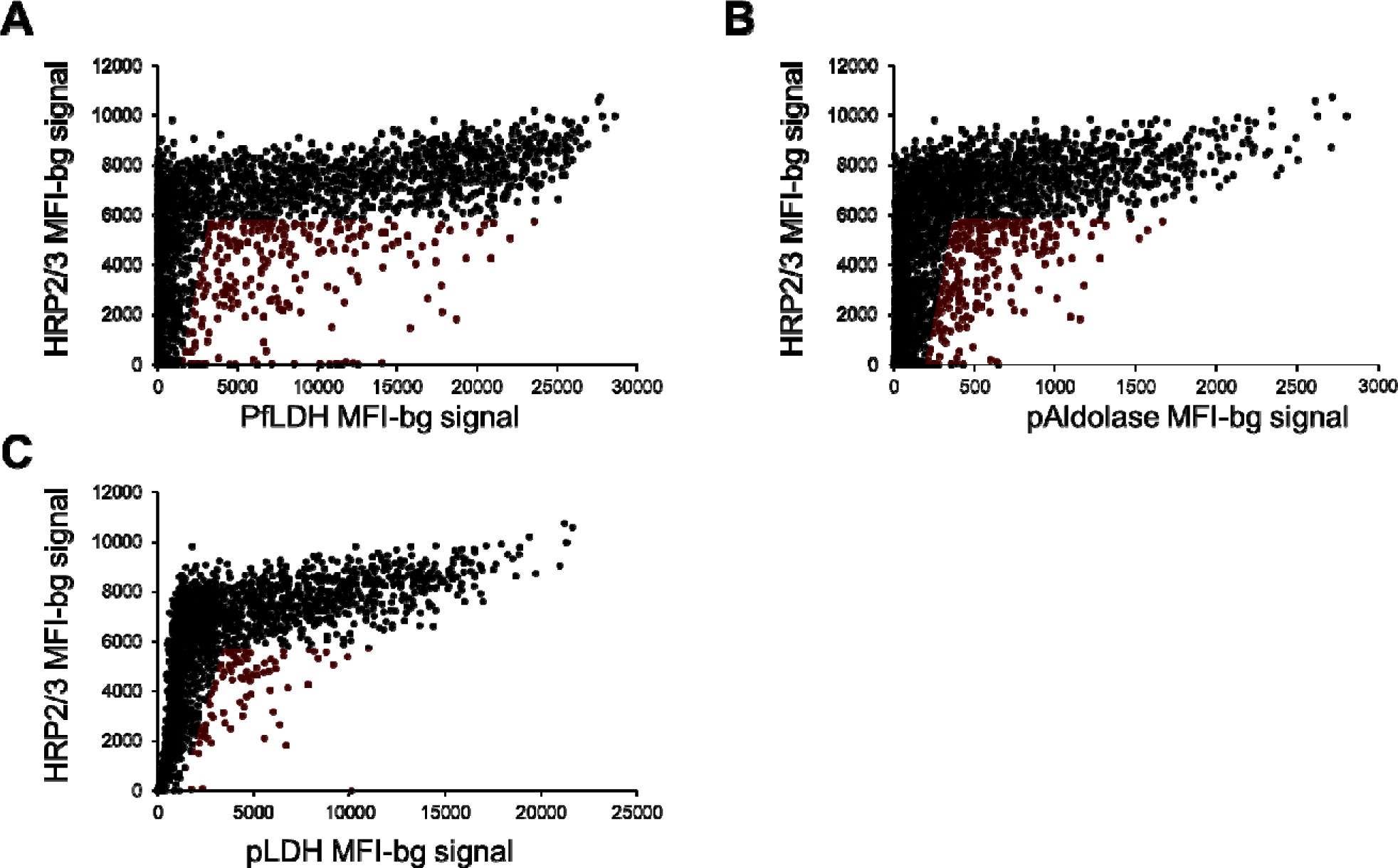
Scatterplots of HRP2 (and HRP3, HRP2/3) antigen levels as compared with other *Plasmodium* antigens. Scatterplots represent data from all 7,847 DBS which were assayed by bead-based multiplex assay for *Plasmodium* antigens. Antigen levels for HRP2/3 versus PfLDH (A), pAldolase (B), and pLDH (C). Red shading indicates DBS with depressed or absent HRP2/3 levels selected for subsequent *pfhrp2* and *pfhrp3* genotyping.

**Figure 3.**
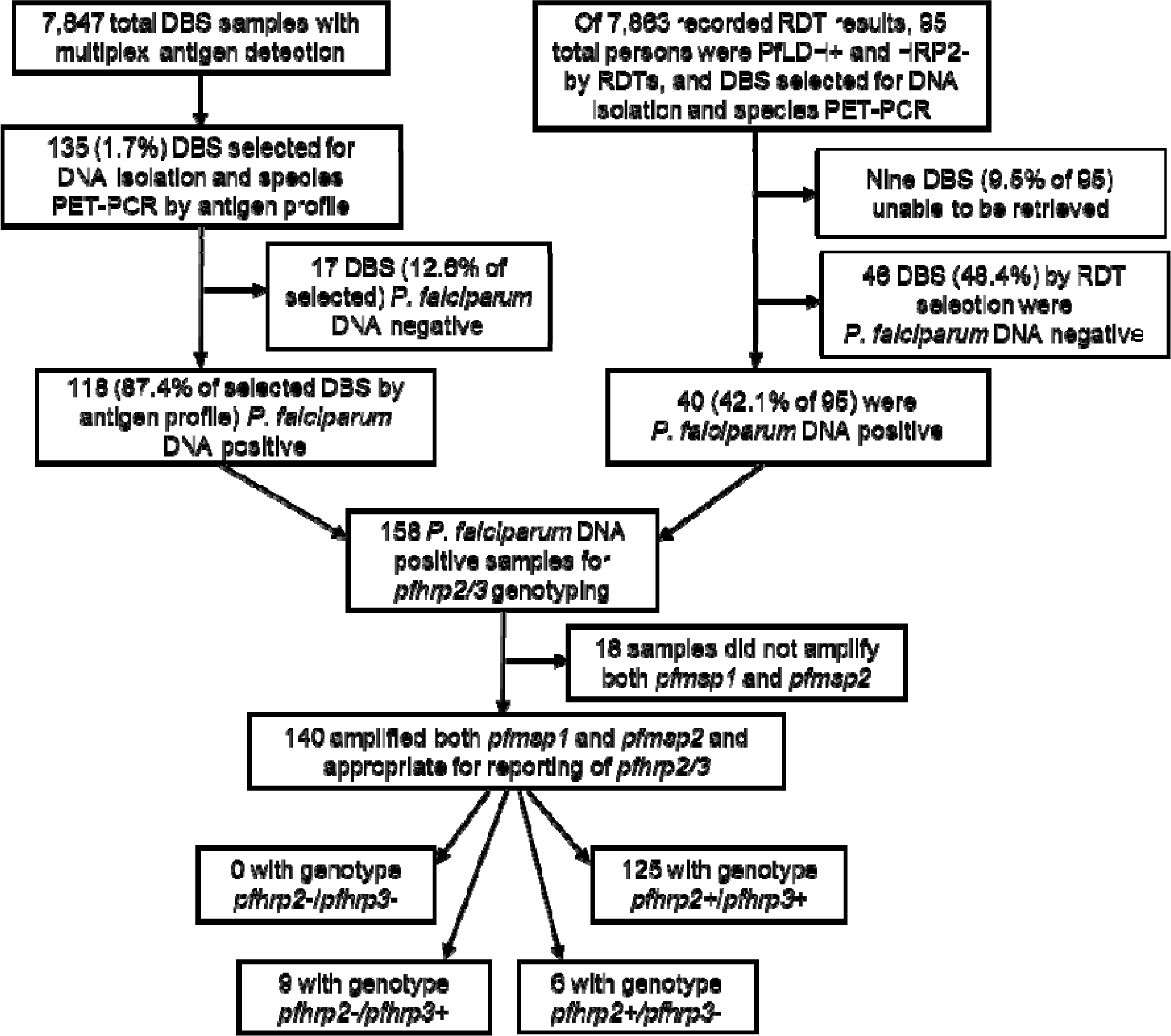
Flow diagram for selection of specimens for genotyping of *pfhrp2* and *pfhrp3* with results: Tanzania, 2021. Selection strategy included consideration of laboratory antigen detection assay (on left), and RDT result at point-of-contact (on right). Terminal boxes display *pfhrp2* and *pfhrp3* deletion genotypes for samples passing all quality steps.

Approximately half (50/95, 52.6%) of the blood samples from these “RDT discordant” persons positive for any *Plasmodium* antigen by the lab multiplex assay, and only 35.8% of these blood samples were positive by both the Pf-pLDH RDT and the Pf-pLDH lab assay.

Between the RDT discordant (n=40) and lab antigen assay (n=118) selection strategies, a total of 137 *P. falciparum* infections were selected to move forward with *pfhrp2/3* genotyping (Figure 3). Most of these *P. falciparum* DNA positive samples (88.6%, 140/158) were also able to amplify the single-copy genes (*pfmsp1* and *pfmsp2* in this study) required for quality assurance reporting of *pfhrp2/3* genotyping results. Of these 140, the majority (89.2%, 125/140) were able to amplify both the *pfhfrp2* and *pfhrp3* genes (a *pfhrp2*+/*pfhrp3*+ genotype), six (4.3%) showed a genotype of *pfhrp2*+/*pfhrp3*-, nine (6.4%) a *pfhrp2-*/*pfhrp3+* genotype, and no parasites from *P. falciparum* infections were found to have the double-deletion *pfhrp2-*/*pfhrp3*- genotype. All nine persons infected with *P. falciparum* with *pfhrp2* deletions were negative on the national RDT for the HRP2 band.

*P. falciparum* infections with single gene deletions for *pfhrp2* or *pfhrp3* were found to be largely dispersed throughout the country, though some clustering was observed in the southern part of Njombe region (Figure 4). Due to the sampling design and rare finding of any deletions, spatial inference was not attempted to denote statistically significant clustering patterns. The nine infections containing *pfhrp2*-deleted *P. falciparum* were from: Dodoma (n=1), Kilimanjaro (n=1), Manyara (n=2) regions in northeast Tanzania, the Njombe region in the south (n=4), and Kagera region in the extreme northwest (n=1). The four *pfhrp2* deletions in Njombe were enrolled at two health facilities with two infections with deleted parasites found in each. From estimates reported in the year 2020 for *P. falciparum* transmission strata by region in Tanzania [26], the majority of the *pfhrp2* deletions (8/9, 88.9%) were found in either “low” or “very low” transmission strata (Supplemental Figure 2). The *pfhrp3*-deleted parasites were found in: Dar es Salam region (n=1), Kagera region (n=2, infections from the same health facility), Njombe region (n=2, infections from different health facilities), and Mbeya region (n=1). The only regions to show parasites both having *pfhrp2* or *pfhrp3* deletions were Kagera and Njombe, and the ones in Njombe were all noted in the far southern area of this region.

**Figure 4.**
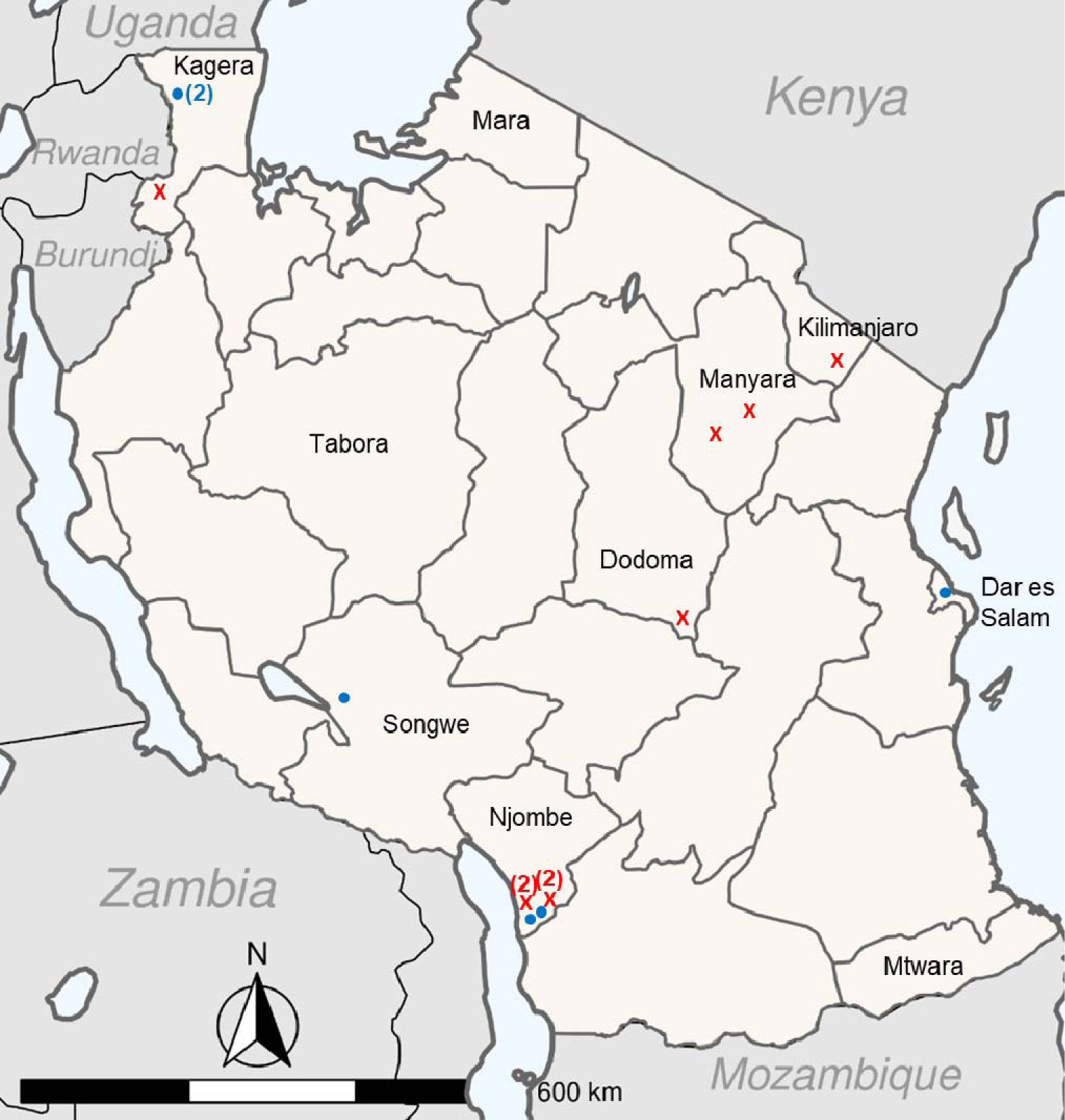
Locations of enrolment sites with identified *P. falciparum* infections with deletions in *pfhrp2* or *pfhrp3*: Tanzania: 2021. Regions that were sampled from are named on the map, with an ‘X’ indicating location of sample identified with *pfhrp2* deletion (n=9) and solid ‘●’ indicating sample identified with *pfhrp3* deletion (n=6). A number in parentheses indicates multiple deletions found at a single site.

## Discussion

Increasing global reports of deletions of the *pfhrp2* (and to lesser extent *pfhrp3*) genes have concerned malaria stakeholders in countries heavily relying on HRP2-based RDTs for clinical diagnosis of *P. falciparum* infection [15, 22]. Enrolling of 7,863 malaria suspected patients in 100 health facilities throughout Tanzania in 2021 who provided a DBS sample found 48.2% of persons with a positive RDT result with very few individuals (n=95, 2.5% of all RDT positives) testing positive for the *P. falciparum* antigen Pf-pLDH, but negative for HRP2.

Importantly, as all persons enrolled in this study were presenting with malaria-like symptoms, from this RDT data alone it could be surmised that *P. falciparum* is the predominant species causing clinical malaria in Tanzania. The concordance in test results between the two independent RDTs shows a low risk of infections with *P. falciparum* not producing high levels of the HRP2/3 antigen(s) – as the phenotypic indicator of deleted or non-functional *pfhrp2* (or *pfhrp3*) genes [9, 13]. In assessing potential *pfhrp2/3* deletions in Tanzania for this survey, multiplex antigen screening including targets for HRP2 and other *Plasmodium* antigens allowed a secondary approach to also compare with RDT results and elucidate *pfhrp2/3* deletions [18, 27]. Of 7,847 DBS assayed by the lab multiplex assay, only 1.7% (n=135) were selected for further molecular assays based on the absence, or very low levels, of HRP2 antigen in the blood sample relative to the other *Plasmodium* targets. The combination of these two approaches (RDT concordance and lab multiplex antigen assay) both pointed to the same phenotypic findings: the vast majority of clinical malaria in Tanzania is being caused by *P. falciparum* producing sufficient HRP2/3 antigen to elicit positive HRP2-based RDT results. The inclusion of *pfhrp2/3* genotyping data for these malaria patients enrolled throughout Tanzania provides further evidence for the very low prevalence of *P. falciparum* parasites causing symptomatic disease that would be missed by routine HRP2-based RDTs currently utilized in Tanzania. When combining the antigen and genotyping data, it is estimated here that 0.24% (95% CI: 0.08-0.39%) of false-negative HRP2-RDTs were due to *pfhrp2* deletions from this survey.

Current recommendations provided by the WHO set a threshold of 5% or greater of clinical *P. falciparum* infections evading HRP2 diagnostics as criteria to re-assess use of HRP2 as a standard diagnostic tool in a setting [17, 21]. For numerous practical reasons, malaria diagnostic strategies are typically made at the national level, so national surveys would need to be inclusive of *P. falciparum* endemic settings throughout a country to most appropriately generate this quantitative estimate and compare to the 5% threshold. This survey design is robust and provides opportunities not only for evaluation of *pfhrp2/3* deletion status in a country, but also assessment of *P. falciparum* parasite diversity (including drug resistance and population genetic markers), estimation of non-falciparum clinical burden, and other malaria clinical comparisons and analyses [6, 28, 29]. From this 2021 Tanzania survey, reports outlining findings for putative drug resistance markers and findings of non-falciparum malaria infections are forthcoming.

Throughout the 100 health facilities where persons were enrolled in this survey, only twelve facilities enrolled persons with *P. falciparum* parasites showing any deletions: seven sites with infecting parasites lacking *pfhrp2*, and five sites with infecting parasites lacking *pfhrp3*.

Furthermore, these twelve sites were broadly spread out throughout the country and inclusive of different sub-populations of *P. falciparum* strains [30]. The notable exception to this generalized finding was the observation of four health facilities in southern Njombe region enrolling persons with *P. falciparum* infections with *pfhrp2* or *pfhrp3* gene deletions. However, even within this “cluster”, of 180 persons enrolled among these three sites, 63% (n=113) were found to have *P. falciparum* infections, meaning that only 3.5% (4/113) of identified *P. falciparum* infections among these three sites showed *pfhrp2* single deletions. Importantly, deletions in the *pfhrp3* gene are not regarded for the WHO 5% *pfhrp2* deletion threshold as this gene product (HRP3) is a truncated form of the HRP2 antigen and is not thought to contribute significantly to the RDT test band intensity when both antigens are present [31]. To this point, all nine *P. falciparum* infections identified in this current Tanzania study which had parasites deleted for *pfhrp2* had produced a negative band result for HRP2 by RDT. Future studies may look to over-sample from this area of the country to see if prevalence and diagnostic impact of *pfhrp2/3* gene deletions is truly higher here versus other regions. The results from this current study are consistent with a previous 2017 Tanzania nationwide survey of border regions which enrolled persons in their households. In this population of persons not actively seeking treatment, 32% of persons were positive to any *Plasmodium* antigen targets, but no *pfhrp2/3* deletions were observed [18]. However, these findings contrast to a 2018 health facility survey enrolling persons in the Kilimanjaro and Tanga regions in the northeast part of the country which estimated *pfhrp2* deletions at 1.6% of *P. falciparum* infections and *pfhrp3* deletions at 52% - though no dual deletions were observed [19]. Different sampling strategies, enrolment during different transmission seasons, and laboratory assays may ultimately lead to differences in prevalence estimates for gene deletions, but results from this current study combined with the previous two studies have consistently shown very low rates of *pfhrp2* deletions.

Though an overall low number of *pfhrp2* deletions were observed in this study, an important finding was that nearly all of these (8/9, 88.9%) were discovered in the relatively lower transmission central and southern regions of the country. This has been observed in previous *pfhrp2/3* deletion studies as well, with these deletion genotypes affecting RDT results relatively more when overall transmission and complexity of infections are reduced [32, 33]. As Tanzania moves towards malaria elimination in the future, reducing overall transmission and the number of *P. falciparum* strains in a patient at any one point in time may increase the risk for deletion genotypes to affect HRP2-RDT accuracy.

This study was subject to limitations which may affect estimates or interpretation of results. Patient enrolment from February to July is mostly inclusive of the rainy season in Tanzania, so prevalence of these deletions leading to false negative HRP2-RDT results during dry season months was not evaluated here [32]. Though the survey was designed to be geographically-representative, persons were only enrolled from 10 of the 26 regions in mainland Tanzania, meaning large areas and *P. falciparum* sub-populations have not been sampled.

*P. falciparum* infections from persons enrolled in 2021 were largely positive for the *pfhrp2/3* genes and produced high levels of the HRP2 antigen to elicit true positive HRP2-RDT results. From evidence generated here, the HRP2 antigen still appears to be a robust tool for identification of *P. falciparum* infection in Tanzania. As these gene-deleted parasites may have a selective advantage over their wild-type counterparts, or potentially be imported, future efforts should continue to monitor for HRP2-RDT validity in the country.

## Data Availability

Data is available upon reasonable request to the authors.

## Acknowledgements

The authors acknowledge the contribution of the following project staff who participated in data collection and/or laboratory processing of samples; Ruth B. Mbwambo, Ramadhani Moshi, Dativa Pereus, Doris Mbata, Ezekiel Malecela, Muhidin Kassim, August Nyaki, Juma Tupa, Idephonce Mathias, Godbless Msaki, Rashid Mtumba, Gasper Lugela, Francis Chambo, Neema Barua, Salome Simba, Hatibu Athumani, Mwanaidi Mtui, Rehema Mtibusa and Tilaus Gustav. The finance, administrative and logistic support team at NIMR: Christopher D. Masaka, Millen A. Meena, Beatrice Mwampeta, Gracia L. Sanga, Neema Manumbu, Halfan Mwanga, Arison Ekoni, Twalipo Mponzi, Pendael Nasary, Denis Byakuzana, Emmanuel Mnzava, Seth Nguhu, Thomas Semdoe and Andrea Kimboi. Management of the National Institute for Medical Research, National Malaria Control Program and Presidents Office-Regional Administration and Local Government (Regional administrative secretaries of the 10 regions, and district officials and staff from all 100 HFs. Technical and logistics support from the Bill and Melinda Gates Foundation team; Philip Welkhoff, Jennifer Gardy, Estee Torok, Sevreina Smith, Janice Culpepper, Lucy Hoffen, Maliha Anwar and Rosalyn Yearly. This study was funded by the Bill and Melinda Gates Foundation (Inv. No. 002202).

## Disclaimer

The findings and conclusions in this report are those of the author(s) and do not necessarily represent the official position of the Centers for Disease Control and Prevention and the Bill and Melinda Gates Foundation.

## Notes

### Competing Interest Statement

The authors have declared no competing interest.

### Author Declarations

Ethics committee of the Medical Research Coordinating Committee (MRCC) of the National Institute for Medical Research (NIMR) gave ethical approval for this work.

## References

1. World Health Organization. World malaria report 2021. Geneva, Switzerland, 2021.

2. Gething PW, Casey DC, Weiss DJ, et al. Mapping Plasmodium falciparum Mortality in Africa between 1990 and 2015. N Engl J Med 2016; 375:2435–45.

3. Keita M, Sogoba N, Kane F, et al. Multiple Resistance Mechanisms to Pyrethroids Insecticides in Anopheles gambiae sensu lato Population From Mali, West Africa. J Infect Dis 2021; 223:S81–S90.

4. Matowo NS, Martin J, Kulkarni MA, et al. An increasing role of pyrethroid-resistant Anopheles funestus in malaria transmission in the Lake Zone, Tanzania. Sci Rep 2021; 11:13457.

4. Berhane A, Anderson K, Mihreteab S, et al. Major Threat to Malaria Control Programs by Plasmodium falciparum Lacking Histidine-Rich Protein 2, Eritrea. Emerg Infect Dis 2018; 24:462–70.

5. Feleke SM, Reichert EN, Mohammed H, et al. Plasmodium falciparum is evolving to escape malaria rapid diagnostic tests in Ethiopia. Nat Microbiol 2021; 6:1289–99.

6. Iriart X, Menard S, Chauvin P, et al. Misdiagnosis of imported falciparum malaria from African areas due to an increased prevalence of pfhrp2/pfhrp3 gene deletion: the Djibouti case. Emerg Microbes Infect 2020; 9:1984–7.

7. Uwimana A, Umulisa N, Venkatesan M, et al. Association of Plasmodium falciparum kelch13 R561H genotypes with delayed parasite clearance in Rwanda: an open-label, single-arm, multicentre, therapeutic efficacy study. Lancet Infect Dis 2021; 21:1120–8.

8. Poti KE, Sullivan DJ, Dondorp AM, Woodrow CJ. HRP2: Transforming Malaria Diagnosis, but with Caveats. Trends Parasitol 2020; 36:112–26.

9. World Health Organizaion. Malaria rapid diagnostic test performance. Results of WHO product testing of malaria RDTs: Round 8 (2016-2018). Geneva, Switzerland, 2018.

10. Plucinski M, Aidoo M, Rogier E. Laboratory Detection of Malaria Antigens: a Strong Tool for Malaria Research, Diagnosis, and Epidemiology. Clin Microbiol Rev 2021; 34:e0025020.

11. Boyce MR, O’Meara WP. Use of malaria RDTs in various health contexts across sub-Saharan Africa: a systematic review. BMC Public Health 2017; 17:470.

12. Gamboa D, Ho MF, Bendezu J, et al. A large proportion of P. falciparum isolates in the Amazon region of Peru lack pfhrp2 and pfhrp3: implications for malaria rapid diagnostic tests. PLoS One 2010; 5:e8091.

14. Akinyi Okoth S, Abdallah JF, Ceron N, et al. Variation in Plasmodium falciparum Histidine-Rich Protein 2 (Pfhrp2) and Plasmodium falciparum Histidine-Rich Protein 3 (Pfhrp3) Gene Deletions in Guyana and Suriname. PLoS One 2015; 10:e0126805.

15. Thomson R, Parr JB, Cheng Q, Chenet S, Perkins M, Cunningham J. Prevalence of Plasmodium falciparum lacking histidine-rich proteins 2 and 3: a systematic review. Bull World Health Organ 2020; 98:558-68F.

16. Cheng Q, Gatton ML, Barnwell J, et al. Plasmodium falciparum parasites lacking histidine-rich protein 2 and 3: a review and recommendations for accurate reporting. Malar J 2014; 13:283.

17. World Health Organization. Response plan to pfhrp2 gene deletions. Geneva, Switzerland, 2019

18. Bakari C, Jones S, Subramaniam G, et al. Community-based surveys for Plasmodium falciparum pfhrp2 and pfhrp3 gene deletions in selected regions of mainland Tanzania. Malar J 2020; 19:391.

19. Kaaya RD, Kavishe RA, Tenu FF, et al. Deletions of the Plasmodium falciparum histidine-rich protein 2/3 genes are common in field isolates from north-eastern Tanzania. Sci Rep 2022; 12:5802.

20. Lyimo BM, Popkin-Hall ZR, Giesbrecht DJ, et al. Potential Opportunities and Challenges of Deploying Next Generation Sequencing and CRISPR-Cas Systems to Support Diagnostics and Surveillance Towards Malaria Control and Elimination in Africa. Front Cell Infect Microbiol 2022; 12:757844.

21. World Health Organization. Master protocol for surveillance of pfhrp2/3 deletions and biobanking to support future research. Geneva, Switzerland, 2020.

22. Rogier E, McCaffery JN, Nace D, et al. Plasmodium falciparum pfhrp2 and pfhrp3 Gene Deletions from Persons with Symptomatic Malaria Infection in Ethiopia, Kenya, Madagascar, and Rwanda. Emerg Infect Dis 2022; 28:608-16.

23. Teyssier NB, Chen A, Duarte EM, Sit R, Greenhouse B, Tessema SK. Optimization of whole-genome sequencing of Plasmodium falciparum from low-density dried blood spot samples. Malar J 2021; 20:116.

24. Jones S, Subramaniam G, Plucinski MM, et al. One-step PCR: A novel protocol for determination of pfhrp2 deletion status in Plasmodium falciparum. PLoS One 2020; 15:e0236369.

25. Rogier E, Bakari C, Mandara CI, et al. Performance of antigen detection for HRP2-based malaria rapid diagnostic tests in community surveys: Tanzania, July-November 2017. Malar J 2022; 21:361.

26. Thawer SG, Chacky F, Runge M, et al. Sub-national stratification of malaria risk in mainland Tanzania: a simplified assembly of survey and routine data. Malar J 2020; 19:177.

27. Plucinski MM, Herman C, Jones S, et al. Screening for Pfhrp2/3-Deleted Plasmodium falciparum, Non-falciparum, and Low-Density Malaria Infections by a Multiplex Antigen Assay. J Infect Dis 2019; 219:437–47.

28. Abuaku B, Amoah LE, Peprah NY, et al. Malaria parasitaemia and mRDT diagnostic performances among symptomatic individuals in selected health care facilities across Ghana. BMC Public Health 2021; 21:239.

29. Amoah LE, Abuaku B, Bukari AH, et al. Contribution of P. falciparum parasites with Pfhrp 2 gene deletions to false negative PfHRP 2 based malaria RDT results in Ghana: A nationwide study of symptomatic malaria patients. PLoS One 2020; 15:e0238749.

30. Moser KA, Madebe RA, Aydemir O, et al. Describing the current status of Plasmodium falciparum population structure and drug resistance within mainland Tanzania using molecular inversion probes. Mol Ecol 2021; 30:100–13.

31. Kong A, Wilson SA, Ah Y, Nace D, Rogier E, Aidoo M. HRP2 and HRP3 cross-reactivity and implications for HRP2-based RDT use in regions with Plasmodium falciparum hrp2 gene deletions. Malar J 2021; 20:207.

32. Kozycki CT, Umulisa N, Rulisa S, et al. False-negative malaria rapid diagnostic tests in Rwanda: impact of Plasmodium falciparum isolates lacking hrp2 and declining malaria transmission. Malar J 2017; 16:123.

33. Molina-de la Fuente I, Benito MJS, Flevaud L, et al. Plasmodium falciparum pfhrp2 and pfhrp3 Gene Deletions in Malaria-Hyperendemic Region, South Sudan. Emerg Infect Dis 2023; 29:154-9.

