## Supplemental Material for "*Plasmodium falciparum pfhrp2* and *pfhrp3* gene deletions among patients enrolled at 100 health facilities throughout Tanzania: February to July 2021"

**Supplemental Information**

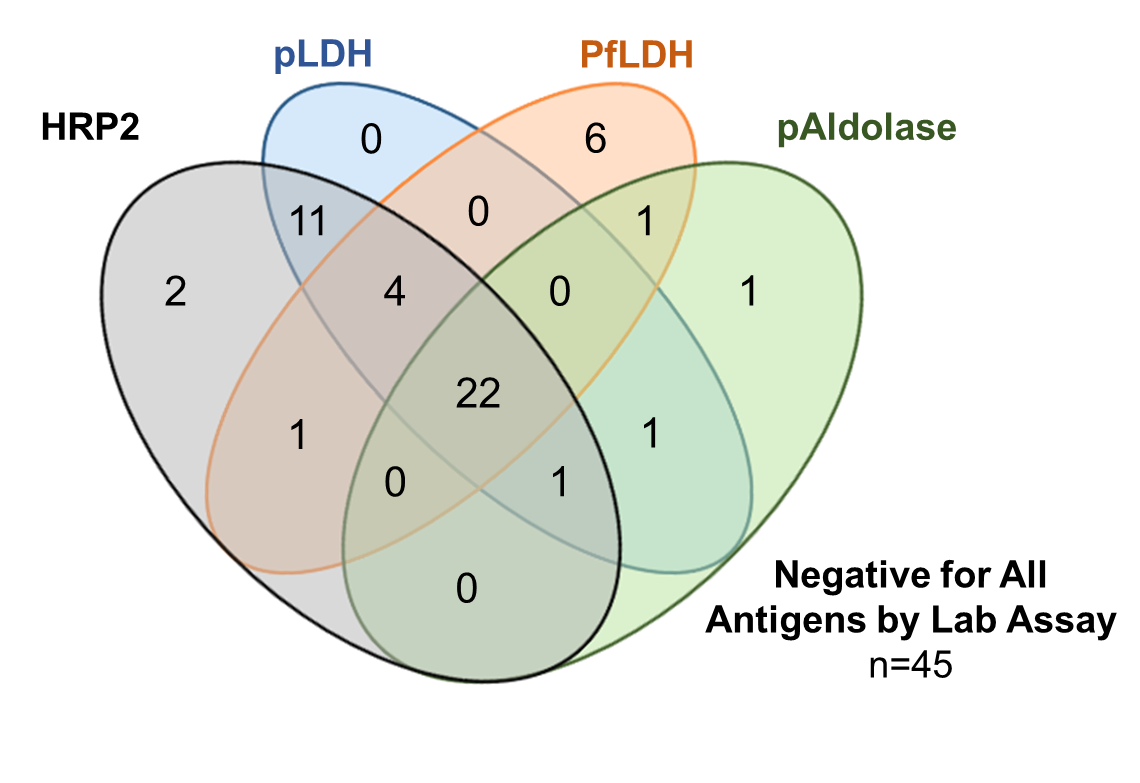

**Supplemental Figure 1**. **Concordance of *Plasmodium* antigen positivity as determined by multiplex lab assay among blood samples for the 95 persons testing RDT discordant: a negative HRP2 band by the national RDT but positive Pf-pLDH band by the research RDT.** Overlapping ovals in Venn diagram show positivity to multiple antigen targets. A total of 50 (52.6%) of these blood samples were positive for at least one antigen by the lab assay, whereas 45 (47.4%) were negative for all *Plasmodium* antigens by lab assay.

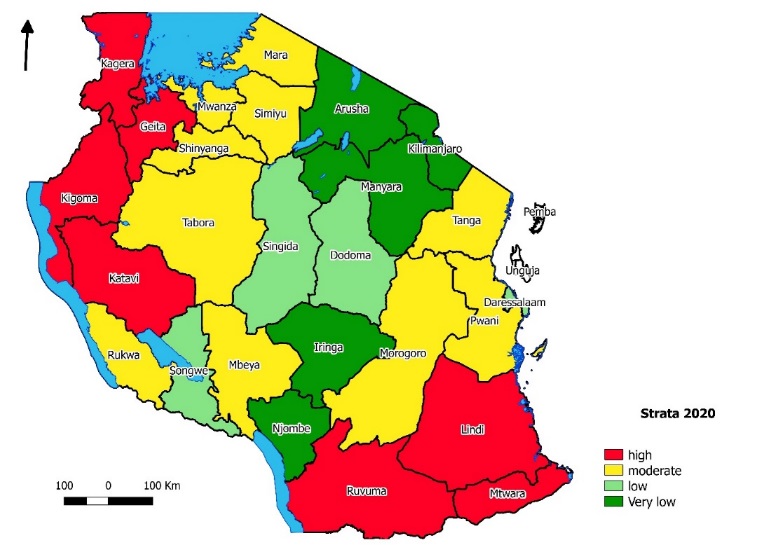

**Supplemental Figure 2. Estimated stratification by region in Tanzania of *P. falciparum* prevalence in school-aged children in Tanzania as estimated by data from surveys and outpatient clinics: 2020.** Authors stratified estimated *P. falciparum* prevalence of five- to 16-year old children into four levels: very low (dark green, <1%), low (light green, 1 to <5%), moderate (yellow, 5 to <30%), and high (red, ≥30%). Full analyses presented in: Twawer, *et al*. Sub-national stratification of malaria risk in mainland Tanzania: a simplified assembly of survey and routine data. *Malaria J* **19**(1): 177 (2020). Scale bar presents distance in kilometers, and directional arrow points north.
